# Neuroligin fragments as blood-based biomarkers for early detection of Alzheimer’s disease

**DOI:** 10.1101/2024.09.05.24313143

**Authors:** Milton Guilherme Forestieri Fernandes, Maxime Pinard, Esen Sokullu, Jean-François Gagnon, Frédéric Calon, Benoit Coulombe, the Consortium for the early identification of Alzheimer’s disease-Quebec (CIMA-Q), Jonathan Brouillette

## Abstract

**INTRODUCTION:** Biomarkers for early detection of Alzheimer’s disease (AD) are essential for improving treatments. Fragments of the synaptic protein neuroligins (NLGNs) are released into the blood due to synaptic degeneration, which occurs in the early stages of AD.

**METHODS:** We used MS2-targeted mass spectrometry on blood samples from the CIMA-Q cohort to assess the potential of NLGN fragments as blood-based biomarkers for amnestic mild cognitive impairment (aMCI), a prodromal stage of AD.

**RESULTS:** We found higher blood levels of certain NLGN fragments in both aMCI and AD patients compared to healthy subjects. Within these same samples, the levels of Tau phosphorylated at various epitopes were higher in AD subjects but not in aMCI individuals.

**DISCUSSION:** Synaptic proteins such as NLGNs could serve as effective biomarkers for detecting the disease in its prodromal stage. This early detection could accelerate diagnosis and therapeutic intervention before neurodegeneration leads to irreversible brain damage.

## 1. BACKGROUND

Early detection of Alzheimer’s disease (AD) is crucial for effective interventions. AD is preceded by two transitional stages: subjective cognitive disorder (SCD), followed by amnestic mild cognitive impairment (aMCI)[1, 2]. However, there is still an urgent need for reliable biomarkers for these stages.

Synapse loss is observed in prodromal AD and correlates with cognitive decline of the patients[3-5]. Neuroligin-1 (NLGN1) is a post-synaptic transmembrane protein almost exclusively localized on glutamatergic synapses and selectively express in the central nervous system (CNS)[6-8]. Previous work has demonstrated that neuronal hyperactivity promotes the cleavage of NLGN1 extracellular domain[9, 10], which is then released into the blood [11, 12]. Recently, our laboratory found that individuals with aMCI have lower levels of NLGN1 in the hippocampal brain region[13]. Also, the level of NLGN1 was found to be decreased even more in AD patients[13].

There is evidence for synaptosomal degeneration in GABAergic inhibitory interneurons in some brain regions of AD patients and animal models[14-17]. Given that the neuroligin isoform NLGN2 is found exclusively at inhibitory synapses, we also aim to evaluate if this other neuroligin isoform is altered in the plasma of aMCI and early AD subjects.

Thus, we hypothesized that neuronal hyperactivity observed in prodromal AD leads to the cleavage of neuroligins (NLGNs), which progressively increases the concentration of soluble NLGN fragments in the plasma of aMCI and AD subjects compared to CTL elderly. Given that blood-based biomarker assays are simple, minimally invasive and cost-effective, measuring plasma level of NLGN fragments could have a direct impact on the development of therapeutics.

To test our hypothesis, we developed an ultra-sensitive technique for quantifying blood levels of NLGN1 and NLGN2. This technique involves Protein Affinity Capture coupled with quantitative Mass Spectrometry (PAC-qMS) to analyze NLGN1 and NLGN2 together in plasma samples from the Consortium for the early identification of Alzheimer’s Disease - Quebec (CIMA-Q) cohort. The demographic characteristics of this cohort are detailed in Supplementary Table 1. We compared the plasma concentrations of NLGN fragments and their potential as biomarkers for aMCI and early AD with those of phosphorylated Tau (pTau) at various epitopes (pTau181, pTau217, and pTau231), which have been identified as promising early biomarkers of AD[18]. The clinical evaluation, inclusion and exclusion criteria, blood sample processing, and method to analyse tau levels for CIMA-Q participants have been previously described in detail[19].

## 2. METHODS

### 2.1 CIMA-Q cohort

The data used in this manuscript were partially obtained from the CIMA-Q cohort, established in 2013 with initial funding from FRQS-Pfizer, a cohort of clinically, cognitively, and neuroimaging-characterized elderly men and women, to collect blood samples with the following goals: (1) to establish an early diagnosis of AD; (2) to make a well-characterized cohort available to the scientific community; (3) to identify new therapeutic targets to prevent or slow cognitive decline in AD; and (4) through subsequent clinical studies. Further details on recruitment methods, exclusion and inclusion criteria, testing materials, and general methodology for the CIMA-Q project can be found in Belleville et *al*.[19]. Diagnostic criteria for AD and aMCI were based on the National Institute on Aging - Alzheimer’s Association and were described in detail in Belleville et *al*.[19].

### 2.2 PAC-qMS sample preparation

5 μg of specific antibody against both NLGN1 (Bio-techne, AF4340) and NLGN2 (Bio-techne, AF5645) was coupled overnight at 37°C to 1 mg of dynabead epoxy M-270 (Thermofisher scientific, Cat #: 14311D) and equilibrated according to the manufacturer’s specifications. Samples and calibration points were prepared by using similar amounts of either patient plasma or commercially availed pooled plasma (Innovative research, Cat #: IPLANAC50ML), respectively. Each sample was diluted in a final volume of 250 μL with 1x PBS (Wisent, Cat #: 311-010-EL). Calibration curves were generated by spiking diluted pooled plasma with known amounts of the recombinant NLGN1 protein (Cedarlane, Cat #: TP307524) and NLGN2 protein (Bio-techne, Cat #: 5645-NL_050) (0, 1, 2, 5, 10, 25, 100 ng). A calibration curve was prepared for each set of 34 tested patient plasma. Equilibrated antibody-coupled magnetic bead was added to the plasma samples (patient or calibration curve points) and NLGN1-NLGN2 capture was performed at 4°C for 1 h on a rotator shaker. Magnetic beads were washed 6 times with fresh HBSEP buffer (10 mM Tris-HCl pH 8.0, 150 mM NaCl, 3 mM EDTA, 0.005% Tween20) and 4 times with water. Captured proteins were eluted with an 33% acetonitrile and 0.4% trifluoroacetic acid elution buffer, dried down by using a speed vacuum centrifuge (Eppendorf) and stored at -70°C until sample digestion. The dried pellets were resuspended in 20 μL of 6 M urea and 100 mM ammonium bicarbonate. 5 μL of reduction buffer (45 mM dithiothreitol, 100 mM ammonium bicarbonate) was added to each sample and incubated at 37°C for 30 min on a thermomixer (Thermofisher scientific) set at 350 RPM. 5 μL of alkylation buffer was added to each tube (100 mM iodoacetamide, 100 mM ammonium bicarbonate) and peptide alkylation was performed at 24°C for 20 min in the dark. Prior to trypsin digestion, 50 μL of 50 mM ammonium bicarbonate was added to reduce the urea concentration under 2 M. Trypsin solution, 5 μL of 100 ng/μL of trypsin sequencing grade from Promega (Promega, Cat #: V5111), 50 mM ammonium bicarbonate was added to each sample. Protein digestion was performed at 37ºC for 18 h. The protein digests were acidified with 9.4 μL formic acid 5% in water and dried with a speed vacuum centrifuge. Prior to LC-MS/MS, protein digests were re-solubilized under agitation for 15 min in 12 μL of 2% ACN-1% formic acid and spiked with heavy-labeled standard peptides specific to NLGN1 and NLGN2 protein forms (JPT Peptide technologies Gmbh, Volmerstrasse, Germany). These peptides were added for signal normalization and retention time confirmation of one NLGN1 peptide (GNYGLLDLIQAL-R*) and one NLGN2 peptide (FPVVNTAYG-R*) where R* is Arg U-^13^C6 and U-^15^N4. Desalting and cleanup of the digests were performed using C18 ZipTip pipette tips (Millipore, Billerica, MA). Eluates were dried down in speed vacuum centrifuge and stored at 20ºC until LC-MS/MS analysis.

### 2.3 Mass spectrometry

Samples were reconstituted under agitation for 15 min in 11 μL of 2% ACN-1% FA and loaded into a 75 μm i.d. × 150 mm Self-Pack C18 column installed in the Easy-nLC II system (Proxeon Biosystems). The buffers used for chromatography were 0.2% formic acid (buffer A) and ACN/0.2% formic acid (buffer B). Peptides were eluted with a two-slope gradient at a flowrate of 250 nL/min. Solvent B first increased from 2 to 35% for 40 min and then from 35 to 85% B for 5 min. The HPLC system was coupled to an Orbitrap Fusion mass spectrometer (Thermo Scientific) through a Nanospray Flex Ion Source. Nanospray and S-lens voltages were set to 1.3-1.8 kV and 60 V, respectively. The capillary temperature was set to 250°C. Targeted MS^2^ spectra were acquired in the Orbitrap with a resolution of 30,000, a target value at 5e4 and a maximum injection time of 150 ms. The peptide fragmentation was performed in the HCD cell at a normalized collision energy of 29%. The peptide m/z and retention time used to detect each peptide are listed in the Supplementary Table 2.

### 2.4 Data analysis

Raw mass spectrometry data was imported in PinPoint (ThermoFisher scientific, Version 1.4.0). Peptide abundance in the calibration curve and each patient sample were calculated using the sum of the area of four transitions per peptides and normalized against the sum of the area of four transition of the heavy-labeled NLGN1 peptide (see Supplementary Table 2). Injected amount (in ng) was used to determine the standard curve linearity for each peptide and to calculate the linear regression equation (see Supplementary Figure 1B). NLGNs amount (in ng) for each sample was evaluated by using the regression equation and the concentration in each sample was than calculated. Lower limit of detection (LLOD) and lower limit of quantification (LLOQ) were set as the 3 and 10 times the background level measured in the lowest amount of the calibration curve.

### 2.5. Statistical analysis

All statistical analyses were executed in R.

#### 2.5.1 Normalization and batch effect correction

After the logarithm transformation (base two) of the blood concentration of the NLGN fragments, values were normalized to eliminate the batch effect, according to the following equation:

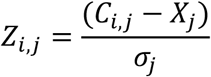

Where:

Z_i, j_ = Z-score of the sample i of the batch j

C_i, j_ = Log_2_ of the blood concentration of the sample i of the batch j

X_j_ = Mean of the log_2_ of the blood concentration of the batch j

σ_j_ = Standard deviation of the log_2_ of the blood concentration of the batch j

#### 2.5.2 Statistical tests

The Shapiro-Wilk test and the Levene’s test were used to check for normality and homoscedasticity. ANOVA, or the Kruskal-Wallis, were performed to assess the differences in multiple groups analysis, and the Tukey’s test for differences between pairs. The number of APOE4 alleles and age of participants were assessed as potential co-factors. A t-test was used to compare the differences in blood concentration of NLGN fragments between sex of participants. The Pearson correlation coefficient along with chi-square were used to evaluate the statistical significance for correlation analysis.

#### 2.5.3 Receiver operating characteristic (ROC) curves and area under the curve (AUC)

Receiver operating characteristic curves were analyzed with the help of the roc() function of the pROC package in R.

## 3. RESULTS

A total of 107 samples from the CIMA-Q dataset were evaluated. Of these, 101 were successfully quantified. The breakdown of these samples by diagnosis is shown in Figure 1A. Figure 1B displays the quantities of samples in which the five fragments of NLGN2 and the single fragment of NLGN1 were quantified. NLGNs were quantified by using standard curves produced with known amounts of each NLGN (Supplementary Figure 1). The sequence of amino acids of these fragments are in Supplementary Table 2, and their positions in both proteins are shown in Supplementary Figure 2. NLGN2 fragments 1 and 2 and the NLGN1 fragment were identified in most of the samples (Figure 1B). The influence of risk factors for AD (APOE4 genes, sex, age and years of education) on the level of NLGN fragments was evaluated (Supplementary Figure 3). Only the presence of two alleles of APOE4 was associated with a higher level of NLGN2 fragment 1 (Supplementary Figure 3A).

**FIGURE 1.**
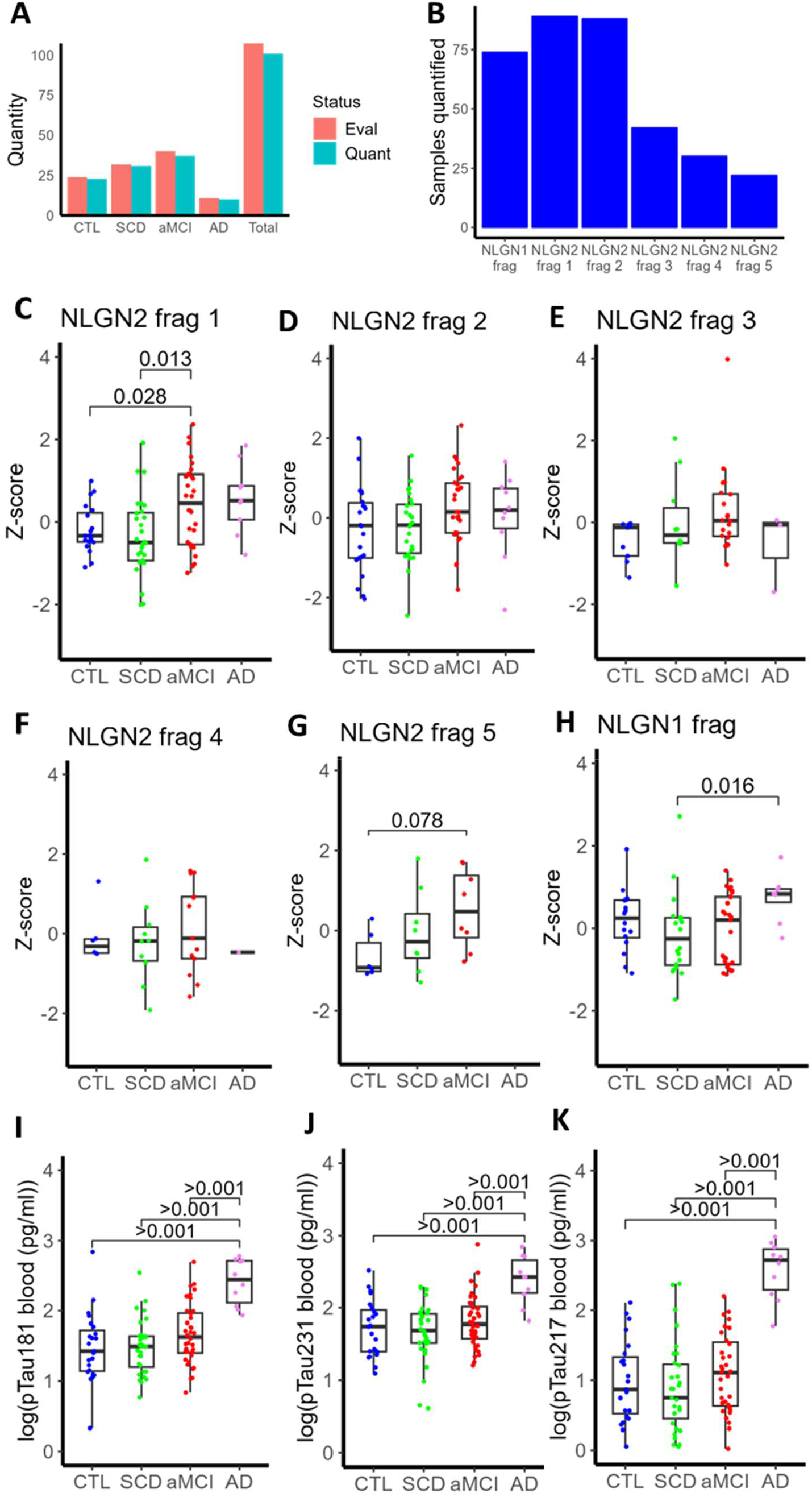
Blood concentration of certain NLGN2 fragments is significantly elevated in amnestic mild cognitive impairment (aMCI). A, Quantity of samples evaluated and quantified by diagnostic group. B, Number of samples in which each peptide was quantified. C**-**H, NLGN fragments levels in blood by diagnostic group. I**-**K, Phosphorylated Tau fragments levels in blood by diagnostic group. Z-score corresponds to the normalized value corrected for batch effect. Significant levels correspond to the p-value calculated by Tukey test. CTL: control, SCD: subjective cognitive disorder, aMCI: amnestic mild cognitive impairment, AD: Alzheimer’s disease. Z-score corresponds to the normalized value corrected for batch effect.

The level of NLGN2 fragment 1 was significantly higher in subjects with aMCI compared to CTL and those with SCD (Figure 1C). The average level of NLGN2 fragment 5 showed a tendency to be higher in aMCI compared to CTL (p-value of 0.078; Figure 1G). The level of NLGN1 fragment was significantly higher in AD patients compared to individuals with SCD (Figure 1H). Additionally, we observed that pTau181, pTau217, and pTau231 were highly effective in differentiating AD patients from CTL, SCD, or aMCI subjects (Figure 1I, J, K). However, none of these pTau biomarkers could distinguish aMCI from CTL or SCD participants, at least in the samples used here to determine blood levels of NLGN1/2. When analyzing a larger number of samples from the CIMA-Q cohort, a significant difference was observed between SCD and aMCI subjects (data not shown). Some past studies have also shown the efficiency of plasma pTau as a biomarker for detecting clinical AD and for distinguishing aMCI from CTL, though with a lower degree of accuracy[20-22]. Because NLGN1/2 fragments can distinguish between different stages of the disease (CTL vs. aMCI, SCD vs. aMCI, and SCD vs. AD), they could be useful for monitoring the progression of the illness.

To further evaluate the potential of NLGN and pTau as biomarkers, we analyzed receiver operating characteristic (ROC) curves and calculated the area under the curve (AUC) for all NLGN fragments and pTau comparing aMCI and AD patients against the control group (Figure 2). For aMCI, NLGN2 fragments 1, 2, 3 and 5 (Figure 2 A, B, C, E) have higher AUC than all pTau (Figure 2 G, H, I). In contrast, when comparing AD to CTL, pTau presented higher AUC values (Fig. 2 G, H, I). We have also observed a significant correlation between the blood level of the NLGN1 fragment, the NLGN2 fragment 1, and all three pTau (Figure 3), indicating that these biomarkers can be used in tandem to better predict AD progression. The correlation between the other NLGN2 fragments and each pTau was not significant (Supplementary Figure 4).

**FIGURE 2.**
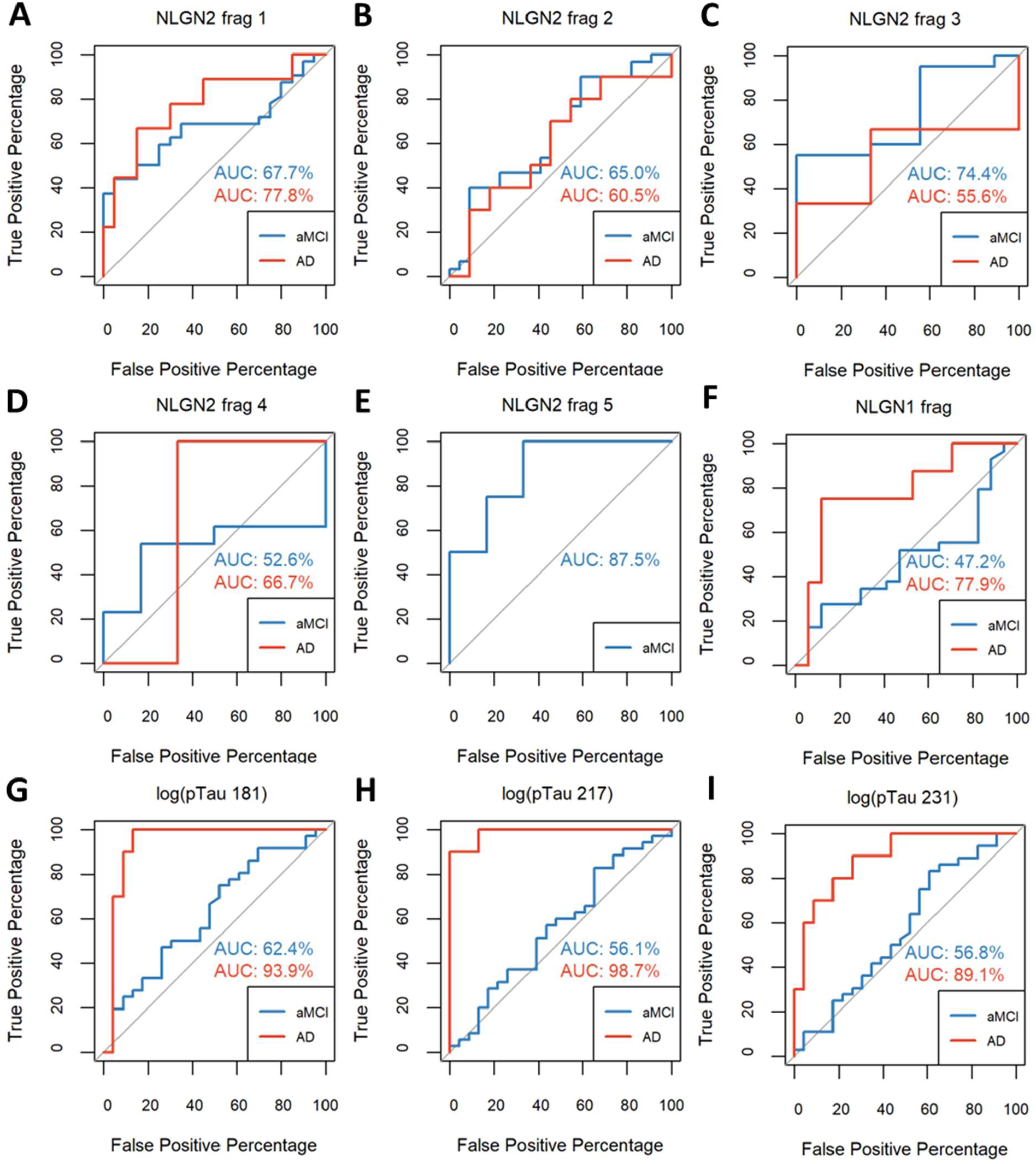
Blood concentration of certain NLGN2 fragments potentially distinguishes amnestic mild cognitive impairment (aMCI). Receiver operating characteristic (ROC) curves with area under the curve (AUC) of NLGN fragments for aMCI and AD in relation to CTL

**FIGURE 3.**
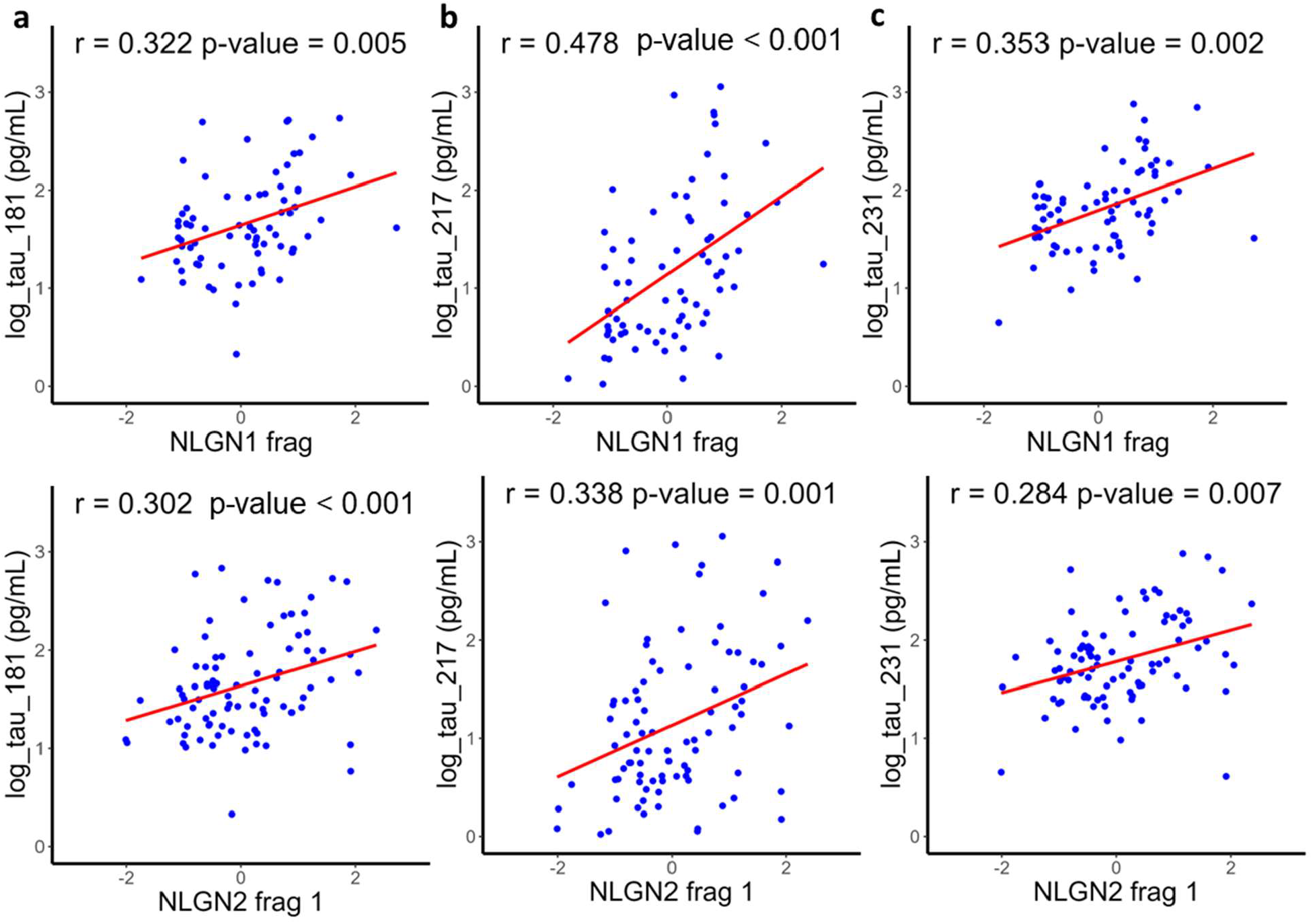
Blood concentration of certain NLGN fragments correlates with blood concentration of phosphorylated Tau. A-C, Correlation between blood concentration of selected fragments of NLGN1 and NLGN2, and blood concentration of phosphorylated Tau. The Pearson correlation coefficient and significant levels are indicated at the top of each plot. The levels of NLGN fragments correspond to the normalized value corrected for batch effect.

To determine if changes in plasma levels of NLGNs are specific for prodromal AD, we used blood samples of individuals in the prodromal stages of neurodegenerative synucleinopathies, including Parkinson’s disease (PD) and dementia with Lewy bodies (DLB). Since the large majority of individuals with REM behaviour disorder (RBD) developed synucleinopathies[23], this condition can be used as a marker of prodromal synucleinopathies. The demographic characteristics of the samples used in this study are provided in Supplementary Table 3. Detailed descriptions of the clinical evaluations, inclusion and exclusion criteria, blood sample processing, and demographics of the Canadian Sleep Research Biobank, from which these samples were obtained, have been previously reported.[24]

We observed no significant difference between RBD patients who developed DLB or PD compared to controls or non-converter (NC) individuals with RBD (Supplementary Figure 5). In most cases, NLGN1 and NLGN2 fragments were under the limit of quantification or not even detected in these participants, suggesting that NLGNs might be a specific marker of prodromal stages of AD.

## 4. DISCUSSION

The development of blood-based biomarkers represents a major breakthrough in improving diagnostic support for AD. In this article, we provide further evidence that blood levels of phosphorylated tau fragments are excellent indicators of AD, as demonstrated in previous studies [18, 22, 25]. We also found that NLGN fragments could be used for detecting the prodromal stage of AD, thus supporting earlier diagnosis and offering more time for therapeutic interventions. Our results are in line with another study showing that Alzheimer patients had lower levels of NLGN1 in plasma exosomes derived from neurons after using an antibody that binds to the extracellular domain of NLGN1.[26] Therefore, using a synaptic protein as a biomarker opens new avenues for early detection of AD, as synaptic degradation precedes widespread tau pathology and neurofibrillary tangles [27-30]. Not only can the use of NLGN fragments as blood-based markers be optimized, but other synaptic proteins involved in synapse degradation also warrant investigation as potential biomarkers for the early stages of AD.

In conclusion, since synapse dysfunction and loss precede neuronal death, identifying changes in synaptic proteins in the blood of individuals with aMCI and early AD would be highly valuable for earlier disease detection and the development of therapeutics while individuals remain autonomous, active, and maintain a good quality of life. In this study, we found that blood levels of certain NLGN1/2 fragments are increased in the prodromal stage of AD. These fragments allow differentiation between various stages of the disease and correlate with pTau181, pTau217, and pTau231. Overall, these results suggest that NLGN fragments could potentially serve as a primary care screening test and be incorporated into a panel of early biomarkers to more accurately track disease progression in clinical practice and therapeutic trials.

## Data Availability

All data produced in the present study are available upon reasonable request to the authors.

## ACKNOWLEDGMENTS

We are grateful to Denis Faubert, Josée Champagne, Sylvain Tessier, and Marguerite Boulos for their assistance in sample processing and mass spectrometric data acquisition. Thank you to Ronald B. Postuma and Nadia Gosselin for providing samples from the Canadian Biobank for Sleep Research.

## CONSENT STATEMENT

All human subjects provided informed consent.

