## Supplementary material for "Neuroligin fragments as blood-based biomarkers for early detection of Alzheimer’s disease"

**Supplementary Table 1: Participants characteristics – CIMA-Q.**

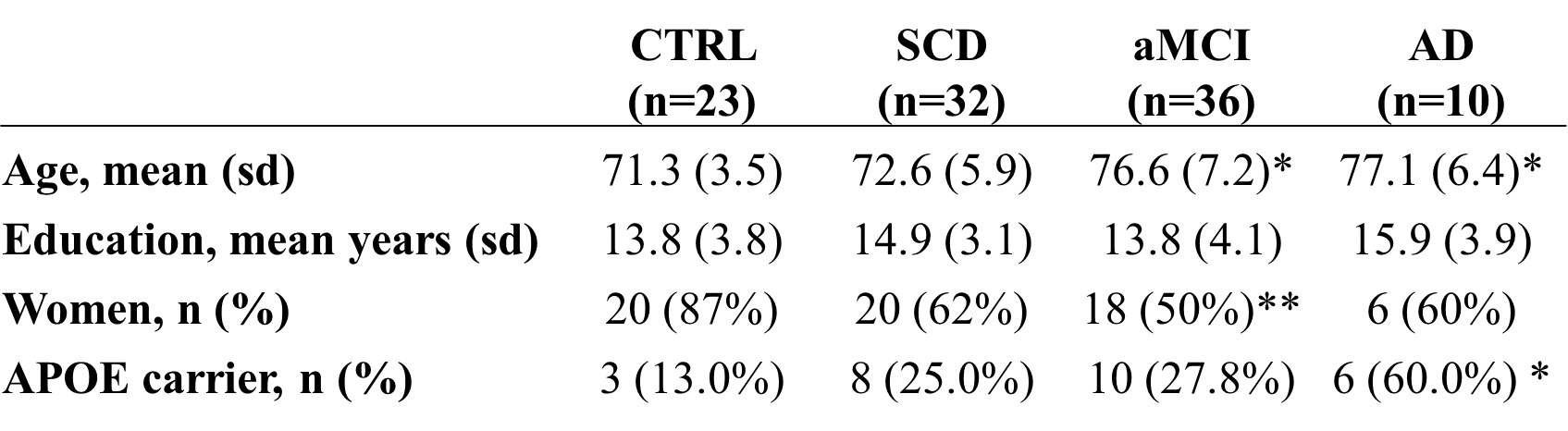

Abbreviations: CTRL – control, SCD – subjective cognitive disorder, aMCI – amnestic mild cognitive impairment, AD - Alzheimer’s disease

**Supplementary Table 2: Peptide sequences and their mass spectrometry characteristics.**

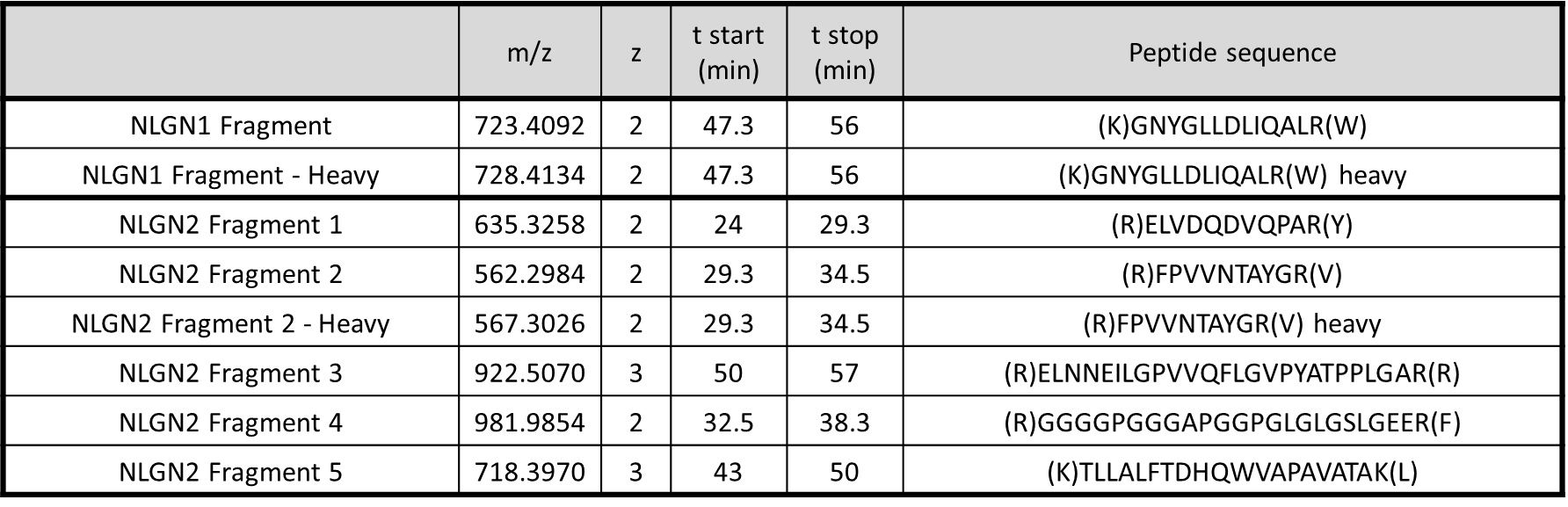

**Supplementary Table 3: Participants characteristics - Canadian Biobank for Sleep Research.**

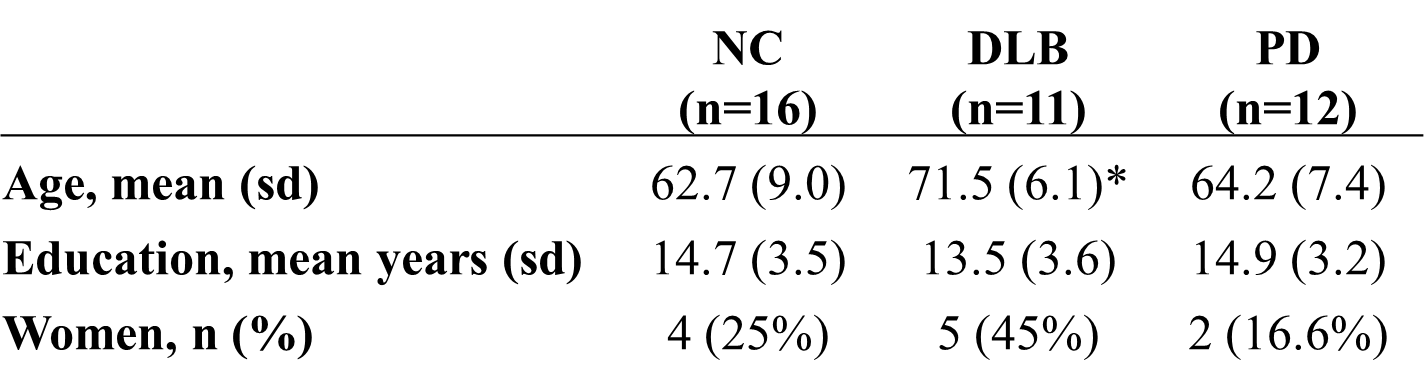

Abbreviations: NC – non-converted, DLB – dementia with Lewy bodies, PD – parkinson’s disease

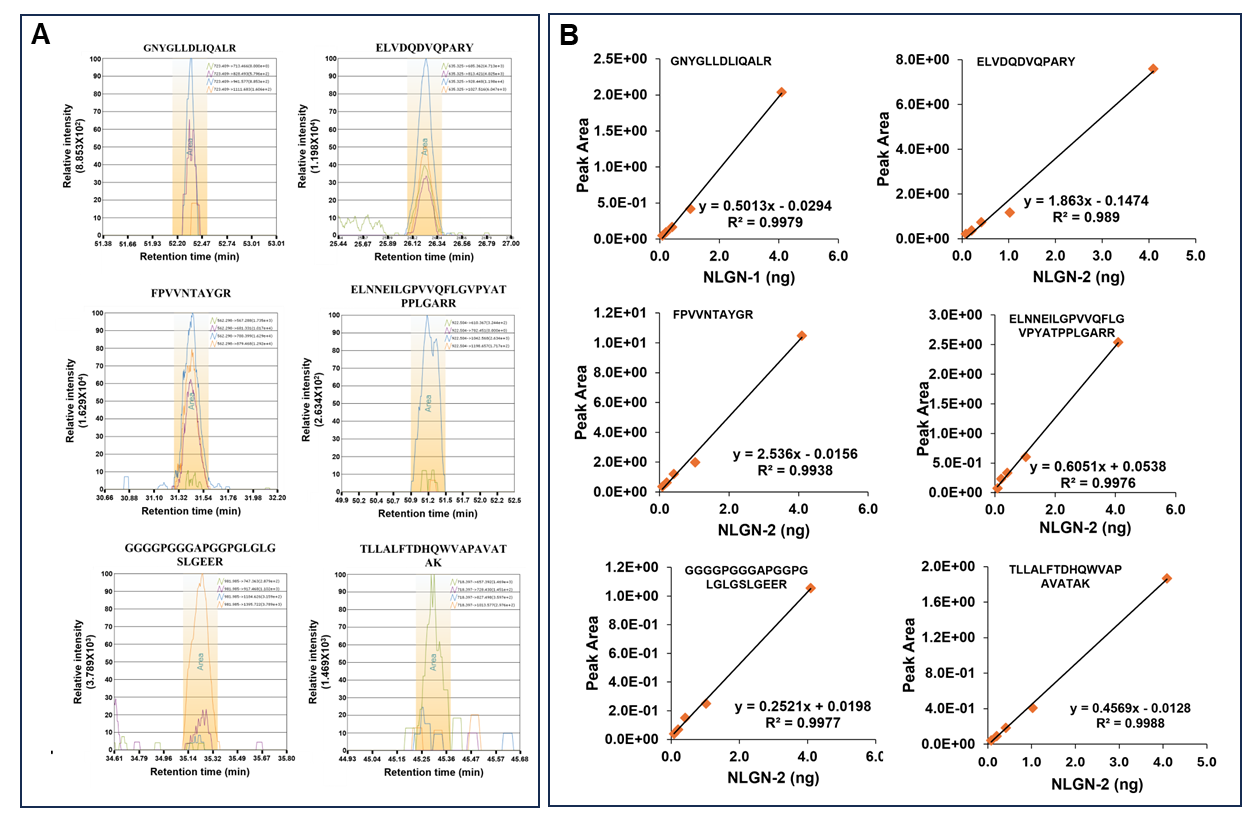

**Supplementary Figure 1:** **NLGNs-enrichment assay chromatograms and standard curves with recombinant NLGNs. a**, Representative chromatograms of each detected peptide in plasma samples, showing endogenous peptide transitions. The x-axis shows elution time in minutes **b**, Representative recombinant NLGNs standard curves for peptides over their linear range. The y-axis peak area represents the value under the curve for each endogenous value relative to NLGN1 heavy isotope-labeled peptide reference (GNYGLLDLIQALR).

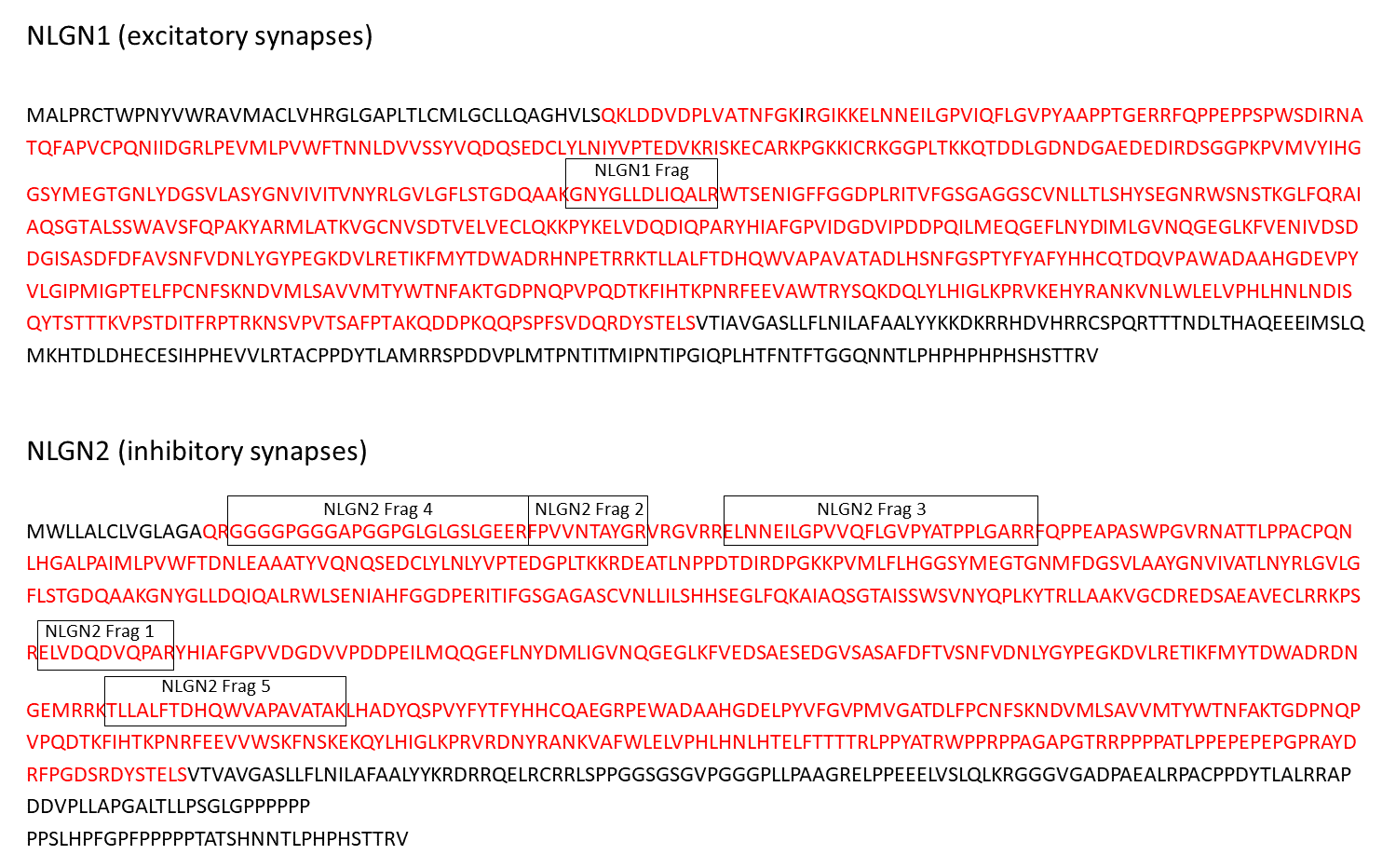

**Supplementary Figure 2: Position of the peptide fragments measured in NLGN1 and NLGN2 amino acid sequence.** The extracellular domain is in red.

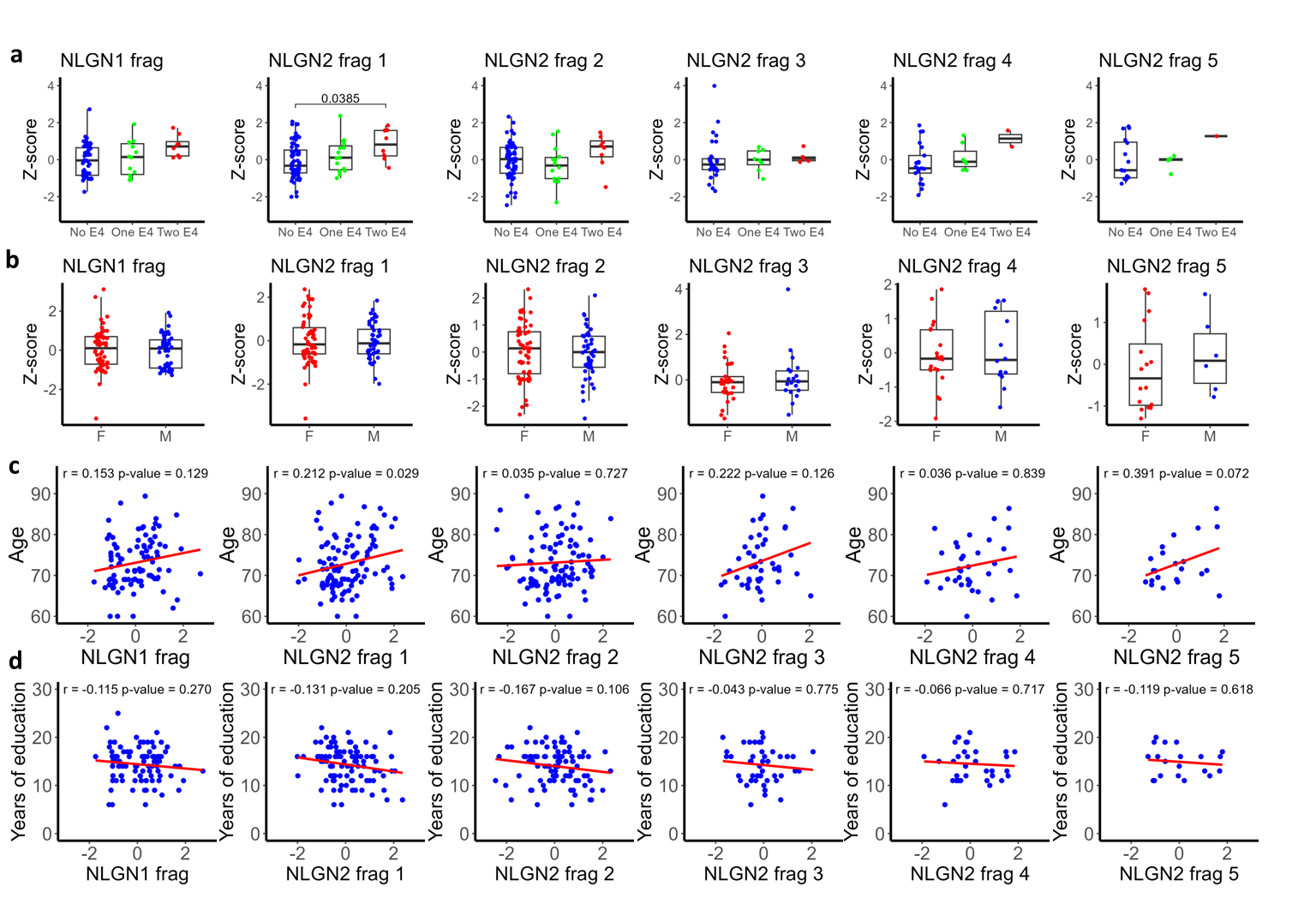

**Supplementary Figure 3:** **Blood concentration of NLGN fragments by APOE4 presence, sex, age and years of education. a, b,** Comparison of NLGN fragments levels in blood by number of APOE4 alleles carried by the subjects sampled **(a)** and by sex **(b)**. Z-score correspond to the normalized value corrected for batch effect, where 0 correspond to the mean and 1 to the standard deviation of the batches in which the indicated fragment was measured, after log2 transformation. **c, d,** Correlation between blood concentration of fragments of NLGN1 and NLGN2, and age (c) and years of education of the subjects sampled. The Pearson correlation coefficient and significant levels are indicated in the top of each plot. The level of NLGN fragments correspond to the normalized value corrected for batch effect, where 0 correspond to the mean and 1 to the standard deviation of the batches in which the indicated fragment was measured, after log2 transformation.

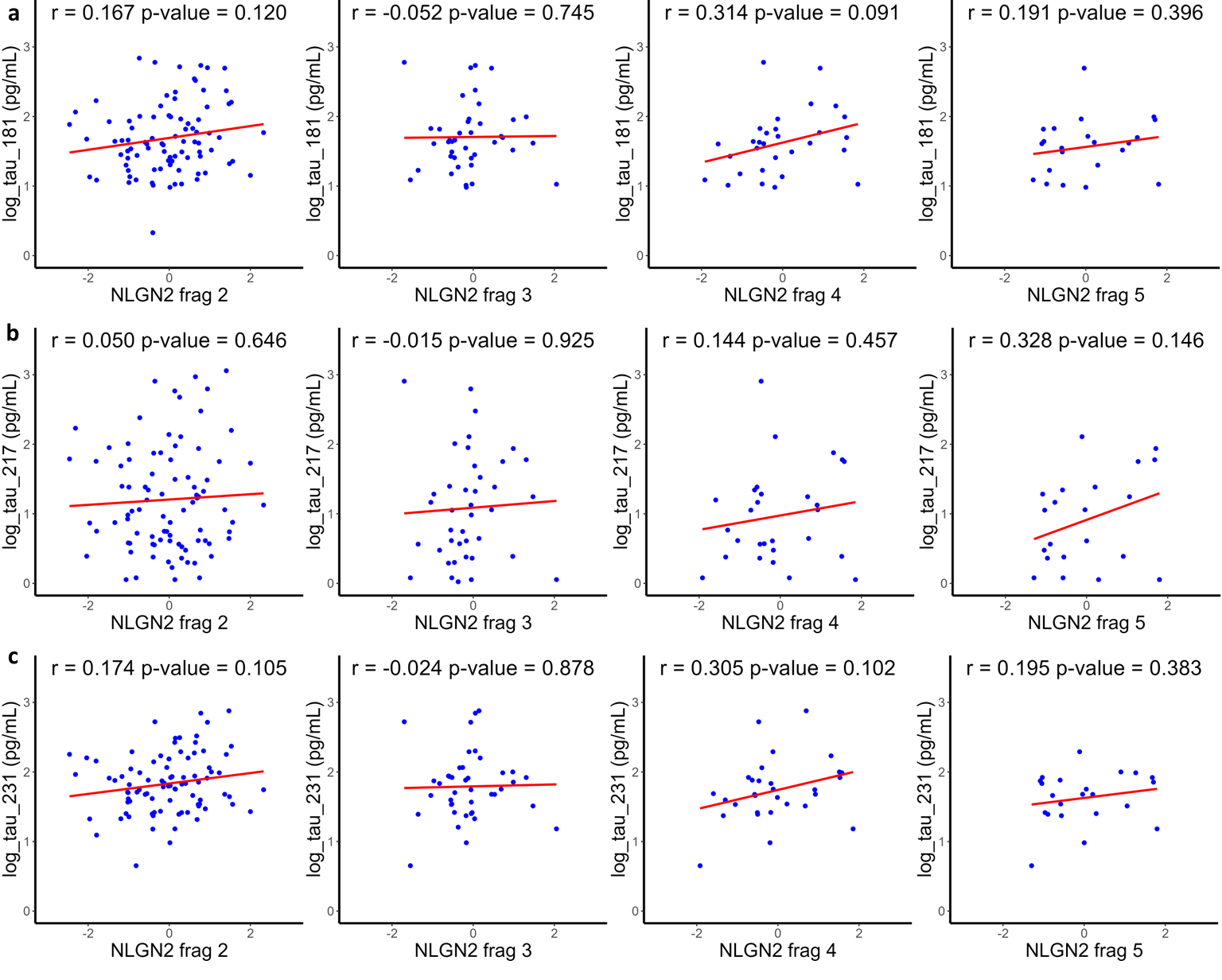

**Supplementary Figure 4:** **Blood concentration of NLGN fragments with blood concentration of phosphorylated Tau (non-significant cases)**. **a-c,** Correlation between blood concentration of fragments of NLGN1 and NLGN2, and blood concentration of Tau phosphorylated at positions 181 (a), 231 (b) and 217 (c). The Pearson correlation coefficient and significant levels are indicated in the top of each plot. The level of NLGN fragments correspond to the normalized value corrected for batch effect, where 0 correspond to the mean and 1 to the standard deviation of the batches in which the indicated fragment was measured, after log2 transformation.

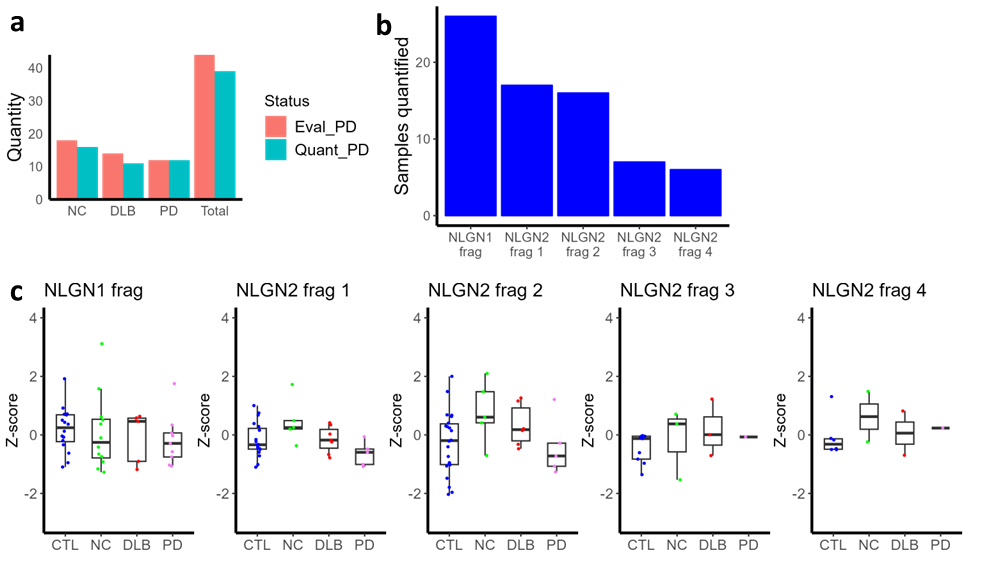

**Supplementary Figure 5: Blood concentration of NLGN fragments in Parkinson disease related diagnoses. a,** Quantity of samples evaluated and quantified by diagnostic group: Non-converted (NC), dementia with Lewy bodies (DLB) and Parkinson’s disease (PD). **b,** Number of samples in which each peptide was quantified (out of 100 total samples for CTL subjects and subjects diagnosed with NC, LBD and PD). **c,** Comparison of NLGN fragments levels in blood of normal subjects (CTL), NC, LBD and PD. Z-score correspond to the normalized value corrected for batch effect, where 0 correspond to the mean and 1 to the standard deviation of the batches in which the indicated fragment was measured, after log2 transformation.
